# Rural Public Health Landscape: Funding, Personnel, Expertise, Community Resources, and Barriers in Rural Settings

**DOI:** 10.1101/2023.06.27.23291769

**Authors:** Olivia Ellison, Vincent Silenzio

## Abstract

**Objectives:** To describe and understand the funding, personnel, expertise, community resources, and issues in rural settings by local public health departments post COVID-19.

**Methods:** Rural county health departments in ten states were sent a survey via web link in the Spring of 2021. 552 responses were collected with a 63% completion rate for all counties surveyed.

**Results:** Most counties utilized public health nurses, administrators, or community health professionals. Of these, 25% had formal education in public health and 10% had public health experience. 65% of respondents disagreed with having adequate funding, staff, and resources. 83% of counties reported working with nonprofits and 43% utilized volunteers. The top two issues in rural public health identified were mental health and substance use.

**Conclusions:** Rural county public health departments do not have the support needed to sustain or advance public health in their specific population.

**Policy implications:** This report gives insight into the needs of rural health in 2021 that can be used to guide policy and funding to support rural health’s specific needs.

## INTRODUCTION

In the United States, approximately 66% of local health departments serve fewer than 50,000 residents.^1^ Even as rural areas make up the majority of local health departments, rural public health is often under studied, departments are expected to provide the same or more services as their urban counterparts, and are underfunded.^1,2^ In a study published in 2002, focusing on three rural states: Alaska, Montana and Wyoming, the authors found that personnel in these public health departments are less likely to have formal training and/or experience^1,3^ and are less likely to be accreditted.^3^ Personnel are more likely to be employed part time and the support of a physician or dentist is used for support or signing death certificates but is also deficient.^1^ Without the support of the state or the community, the rural public health department is set up for potential failure.^1^

In terms of infrastructure, rural public health departments customarily serve smaller communities, leading to smaller tax bases to support public health activities. Charity demand is also higher in these areas due to lower insurance coverage rates, which contributes to having fewer resources available to support epidemiologists, physicians, etc.^4^ Even when considering federal policies targeting underserved areas, they can often unintentionally benefit wealthier communities. Examples of these programs are the Medicare Hospital Readmissions Reduction Program and the federal 340B Drug Discount Program. Additionally, urban areas have seen sizable advances with help of federal programs like the Accountable Health Communities Model and the Prevention of Public Health Fund, which have yet to translate considerably to rural communities.^4^

A rural county can be defined as having a small population, restricted access to resources, and lower density, unlike more urban areas.^3^ Rural-urban health disparities have continued to widen, since even before the COVID-19 pandemic. According to *Healthy People 2020*, between 2007 to 2017 the mortality disparity increased for 5 of the 7 major causes of death: cancer, heart disease, suicide, diabetes, and chronic obstructive pulmonary disease.^4,5^ Data also suggests that those most vulnerable to the spread of disease by injecting drugs are those in rural areas. They traditionally have fewer preventative measures, such as immunizations, sexual education, and tobacco control.^4,6^ Maternal and child health also suffers at the hands of underfunding as morbidity and mortality rates continue to rise after 100 years of decline.^3^ The mothers most at risk of severe adverse outcomes are Black women in the rural South. Rural areas are plagued by racism which has exacerbated the public health crises occurring in rural populations and is confounded by racism not being considered a factor in public health outcomes.^3^

An underutilized resource for rural health communities are community healthcare workers (CHWs), and studies have their shown tremendous opportunities to create better health outcomes in rural communities.^7^ A recent literature review focused on rural CHWs impact reported finding virtually all studies had positive outcomes and a positive return on investment in studies focused on cost. This is significant because this review provides support for community engagement to improve access to care and the cost effectiveness.^7^ Barriers to rural health departments implementing a CHW program may include the lack of standardized training, funding, limited access to transportation, technology, cultural barriers, poor healthcare system infrastructure, sustainability, and the recommendation from the U.S. Department of Health and Human Services, Health Resources and Services Administration to conduct a comprehensive community health needs assessment before identifying a type of CHW program to implement.^8^

In six rural states studied in 2009, when asked about their funding from the Centers for Disease Control, respondents disclosed that the majority of funding is retained at the state level for statewide initiatives.^9^ The study found that there was not enough local funding to address major issues including, cancer control, injury prevention or diabetes. Also, to note, federal funding commonly has a restriction on how much money can be used for administrative costs which is easily used up by the state before trickling down to the local health departments. The money that remains is usually distributed by a “mini-grant” process and does not always represent adequate amounts needed to start programs let alone sustain them.^9^ This is also assuming that a local health department has the expertise or staffing to identify and apply for these grants. Occasionally, a health department must turn down a grant because of lack of staffing to create or sustain a program. In support of this barrier, there is the perception that a program may not necessarily be qualified for a grant based on their ability to collect data and prove effectiveness through evidence-based programming, or the inability to reach their audience due to the lack of public transportation.^9^

## METHODS

The definition of *rural* used in this paper is based on that used by the United States Census and the Office of Management and Budget. The United States Census considers any area not urban, urbanized areas of 50,000 or more people, or urban clusters of 2,500 – 49,999 people, as rural. The Office of Management and Budget considers counties of less than 50,000 people as rural. ^10^ Counties with less than 50,000 people were identified for inclusion using the census data, *Population Data by County 2010 Census Data with* Population Estimates. All counties with more than 50,000 people were excluded from the 2010 census or the *2020 estimate*. ^11^ The results of the 2010 census were utilized due to the delays in the 2020 census caused by the COVID-19 pandemic. Counties were then filtered by size and those that had fewer than 50,000 people were calculated and arranged by state.

| State | # Of Rural Counties | State | # Of Rural Counties | State | # Of Rural Counties |
| --- | --- | --- | --- | --- | --- |
| Hawaii | 1 | California | 15 | Minnesota | 68 |
| Texas | 185 | Kansas | 95 | Iowa | 88 |
| Nebraska | 88 | Mississippi | 68 | North Carolina | 47 |
| Montana | 50 | Oregon | 18 | Illinois | 73 |
| Alaska | 26 | Georgia | 118 | Louisiana | 43 |
| New Mexico | 22 | Missouri | 93 | Wisconsin | 43 |
| North Dakota | 49 | Michigan | 48 | Pennsylvania | 23 |
| Colorado | 49 | Kentucky | 101 | New York | 17 |
| Idaho | 38 | Oklahoma | 63 | Tennessee | 63 |
| Nevada | 12 | Virginia | 96 | Arkansas | 62 |
| South Dakota | 63 | Wyoming | 21 | West Virginia | 44 |
| Utah | 20 | Washington | 18 | Indiana | 65 |
| Vermont | 10 | South Carolina | 22 | Arizona | 4 |
| Alabama | 40 | Florida | 26 | Massachusetts | 2 |
| Ohio | 39 | Maryland | 6 | South Carolina | 22 |
| Maine | 7 | New Hampshire | 3 | Rhode Island | 1 |

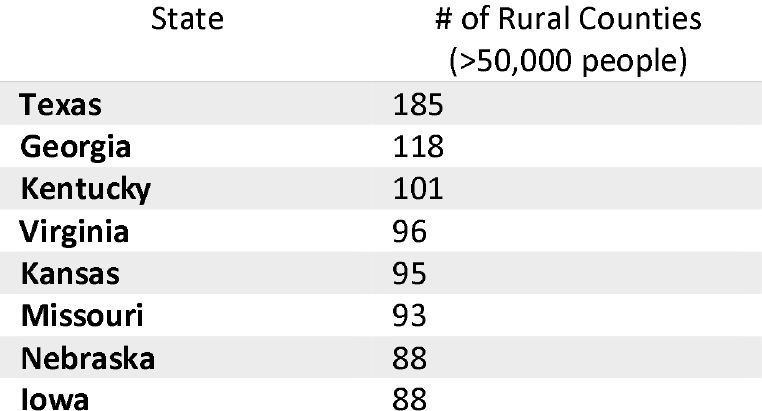
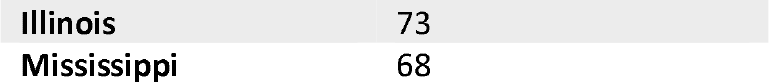

Ten states were identified having the highest number of rural counties in the United States. Texas had the highest number of rural counties with 185, and Mississippi had the lowest with 68 rural counties.

State websites were utilized to find contact information for each of health departments in the identified rural counties. Every county did not have a designated health department or a designated person responsible for the health of the county. In the state of Texas, only 118 of the 185 rural counties had health departments, the remaining counties were served under the umbrella of the state and provided no overlapping services locally. The Texas Department of State Health Services only had contact information for 34 of the 118 (18%) rural defined county’s health departments. Overall, we found 88.7% of contact information publicly available for counties/districts. We also encouraged district representatives to forward on to their county health departments within their district if individual county information was not publicly available. In total, 829 counties were contacted by county or district contact. We solicited help through rural public health associations, National Association of County and City Health Officials (NACCHO), LinkedIn, local state public health associations affiliated with the American Public Health Association (APHA), regional, district, and state offices through their rural, local, and/or communications departments and individual health department websites.

Some counties collaborated with other jurisdictions and shared personnel, so their district representatives were asked to fill out the survey. Those personnel were divided among the number of rural counties they serve. The data may be affected or influenced by a district having a non-rural county within but will still be a valid representation for how what resources rural counties have access to because how often that occurs. For example, some counties share public health resources with various local counties and are represented by a single district umbrella.

### Survey

The survey was designed specifically for this project due to lack of consistent instruments used in public health personnel surveys. Surveys that were referenced in the article, *Rural–Urban Differences in the Public Health Workforce: Local Health Departments in 3 Rural Western* States by Roger A. Rosenblatt, MD, MPH, Susan Casey, PhD, and Mary Richardson, PhD, MHA, were not available or did not have enough information to design a survey based on their initial research on public health staffing. The survey specifically asked which “rural” public health departments the participant answering served due to some health departments serving more than one rural health department. Therefore, the pool of resources would be averaged across the # of rural departments that particular health department serves which excludes non-rural departments they may also serve. The survey included items pertaining to demographics, type of public health personnel, utilization of volunteers and nonprofit relationships pre and post COVID-19, types of funding, perception of funding, resources, expertise, and overall tools to be successful and top ten issues plaguing their jurisdiction in no specific order.

## RESULTS

Survey responses were excluded if the respondent did not consent to the survey, if the population size selected was more than 50,000 per the rural county definition with no rural counties selected, if no state and county were selected or if the answer was duplicative for the specific county being served, or if the survey was not completed further than identifying the state and counties served. Duplicative answers were differentiated by if the respondent served more than one county. The answer that included more than one county was given priority to keep the consistency and reliability of the data. If there was no difference on how many counties were served by the survey answer, the response was chosen at random. The surveys counted were those considered finished which included questions left intentionally blank. If more than one county was served, the quantitative answers were divided by the number of rural counties selected.

### Descriptive Statistics

A total of 552 responses were collected, 29.2% were incomplete (n=161), 0.01% were marked complete but had no data (n=4), 0.007% were not states/counties being studied (n=4), 0.005% had no consent (n=3), 0.005% had no counties selected (n=3), 12.1% were duplicate answers (n=67), and 43.8% were complete responses (n=242). On average, over the 10 states surveyed when counted by county, we had a completion rate of 63%. The lowest response was from the state of Iowa at 36% and the highest being Mississippi at 100%. Mississippi’s response was from one state official instead of individual county. Three counties were served by multiple health departments: in Georgia the county of Dooly and Macon, and in Texas the county of Borden. Missouri had the highest number of individual responses (n=54) and Mississippi had the least (n=1). Respondents were allowed to choose more than one size of population served, 60% responded counties outside of metro or micro areas (n=151), 37% responded counties described as micro area (10,000-49,999 people) (n=93), and 2% being a metro area (<50,000 people) (n=6).

### Personnel

For full time personnel weighted by counties served, counties reported having less than one physician, physician assistant, nurse practitioner, nutritionist, health educator, social worker, community healthcare worker, or volunteer. The counties on average had 3.2 nurses, 1.1 management positions, 1.9 administrators, and 1.5 other direct care providers. Of these full-time personnel, 25% of them had education in public health and 10% had previous experience in public health. For part time personnel weighted by counties served, all positions were reported as having less than one person or position. Of those that did serve in part time roles, 26% had education in public health and 16% had previous experience in public health.

### Resources

For all 10 states, the state general fund was ranked the highest source of revenue for rural counties at 24%, the second highest ranked was tied for federal revenue streams and the state general fund at 27% respectively. The third highest ranked was Medicare/Medicaid at 30% then grants at 31%. This question collected 208 responses with the last ranking being left blank intentionally twice for 206 responses.

When asked about how adequate resources were, 237 responses were collected. 65% of the respondents reported “Somewhat Disagree” or “Strongly Disagree” on having adequate funding. 62% reported “Somewhat Disagree” or “Strongly Disagree” on having adequate staff to serve their community. 58% of respondents reported “Somewhat Agree” or “Strongly Agree” that they had adequate expertise in their department. When asked overall if they had enough resources including funding, staff, and expertise, 66% “Somewhat Disagree” or “Strongly Disagree”.

Of 237 responses, 83% had nonprofit partnerships (n=196). Of those with partnerships, 39% said yes COVID-19 effected these partnerships, 42% responded COVID-19 did not affect these partnerships and 18% responded “Maybe”. The types of nonprofits included local government programming (state and county), community boards and clubs, churches, clinics, hospitals, federally qualified health centers, schools, universities, nursing homes, and food pantries. When asked about utilizing volunteers, only 43% of the counties surveyed worked with volunteers (n=102). Of those who answered “Yes”, 49% responded that COVID-19 effected their ability to work with volunteers, 36% said COVID-19 did not, and 15% said “Maybe”. As for community interest, from 239 responses, 79% answered “Probably Yes” or “Definitely Yes” when asked if they had interest in their community to improve health outcomes.

The two top issues identified from the list of Healthy People 2020 topics plaguing rural communities were “Mental Health and Mental Disorders” and “Substance Abuse” at (n=170). Additionally, “Diabetes” (n=169), “Access to Health Services” (n=155), and “Heart Disease and Stroke” (n=149) were identified as prevalent issues health departments were working to improve outcomes of.

When asked to elaborate on what other resources these rural health departments may benefit from the common themes included: funding issues lack thereof and funding being tied to specific issues or populations (n=34), a need for public health personnel and infrastructure (n=21), transportation concerns (n=14), a need for mental health/behavioral health services (n=13), an interest in clinics specifically for low income, dental and eye services (n=9), and access (n=9).

**Table 1.** Rate how adequate each of these are funding, staff, expertise and overall.

| Variable | Strongly Agree<br>(n=241) |  | Somewhat<br>Agree (n=241) |  | Neither Agree<br>nor Disagree<br>(n=241) |  | Somewhat<br>Disagree<br>(n=241) |  | Strongly<br>Disagree<br>(n=241) |  |
| --- | --- | --- | --- | --- | --- | --- | --- | --- | --- | --- |
| **Funding** | 7 | 3% | 50 | 21% | 29 | 12% | 89 | 38% | 63 | 27% |
| **Staff** | 8 | 3% | 58 | 24% | 25 | 11% | 91 | 38% | 56 | 24% |
| **Expertise** | 43 | 18% | 95 | 40% | 39 | 16% | 52 | 22% | 9 | 4% |
| **Overall** | 4 | 2% | 38 | 16% | 38 | 16% | 98 | 41% | 60 | 25% |

## DISCUSSION

When weighted by numbers of counties served, full time and part time, public health nurses were the most utilized type of personnel then followed administrators and other direct care supervisors. This leaves ample opportunity for other types of public health professionals to contribute to rural public health. Only 25% of the full time rural public health workforce surveyed had public health education and only 10% had professional public health experience. Of these professionals, physicians, health educators, social workers, community health workers and nurse practitioners were <1 per county. Comparatively, when surveyed on the adequacy of their infrastructure, specifically their departments expertise, 58% strongly agreed or somewhat agreed that the department possessed adequate expertise.

This identifies a gap in hiring capability or availability of public health graduates to work in these departments. This is supported by 62% of survey respondents answering that they strongly or somewhat disagreed that they had adequate staffing. To combat this, more funding would attract new candidates, as well as internships, work flexibility, and marketing of opportunities. A variety of type of staff and utilization of potentially unconventional or dual-purpose public health professionals like nutritionists could be beneficial.

Community connection is an underutilized way to support public health infrastructure. This is supported by <1 full time or part volunteer in the health departments surveyed but health departments identifying a 79% *probability of probably yes* or definitely yes in community interest. Although COVID-19 has affected their ability to work with volunteers by 49%, as the pandemic becomes more endemic, the opportunity arises. Mobilizing community healthcare workers as volunteers could be a solution to cycle of lack of funding contributing to lack of staffing. With ample community interest, a community can train volunteers to start conversations with their neighbors to promote positive health outcomes.

When asked about types of nonprofits counties worked with, responses included an extensive list: substance use, domestic violence shelters, Red Cross, United Way, Arc, Salvation Army, churches, schools, colleges, federal, state and local programs, community boards and organizations, food banks, mental health providers, clinics, community healthcare providers, etc. This further supports how vital community connection and contribution may be to filling in the gaps left by the lack of funding and staffing. This also illustrates what types of non-traditional resources can be tapped into to benefit the health outcomes of a community. These areas may warrant further study as types of resources rural communities are tapping into to identify not previously known issues in the rural community.

The types of funding for public health departments varied. Federal revenue streams comprised the majority at 24%, followed by the state, then Medicare/Medicaid, then fees and fines, and grants. When stratified by state, only 4 of the 10 states surveyed reported federal funding as their highest revenue stream. Illinois was included in this count but tied for their top revenue stream was grant money. Kansas also listed grants as their highest revenue stream. This illuminates either Illinois and Kansas’ investment into grant writing or the inaccessibility of grants to other states. This topic could be furthered explored as to why other states do not or cannot take advantage of grant opportunities. Also, to note of importance, not all states chose to not expand immediately or have not expanded their Medicaid/Medicare programs under the Affordable Care Act implemented by the Obama administration.

As previously mentioned, survey respondents reported inadequate funding (65%) and inadequate staffing (62%) which contributed to inadequate resources and infrastructure overall (66%). This was supported by the qualitative answers categorized by topic with 22% answering funding/funding issues and 14% answering personnel/infrastructure as resources their department would benefit from. Funding issues included funding being tied to specific issues or populations. This contradicts studies done two decades ago that identified expertise as being a major contributor to lack of success in rural health departments. Funders and policymakers must consider whether their parameters for funding are benefitting specific populations enough with targeting funding or if they are contributing to a larger issue of lack of funding for health departments to improve health outcomes for more people.

## Limitations

Limitations of this study include the potential of uneven weighting for county resources. When asked which counties the jurisdictional health department served, only rural counties were given as options not metro counties. Health departments may have also served urban counties which may overestimate the number of resources the rural counties they serve are provided. At the time of publication some jurisdictions may still have been benefitting from COVID-19 emergency funding which could have inflated their answers on resources, funding and expertise. In the survey, the only question required to answer was the size of population. The results may be biased based on how many surveys were completed in full or had intentionally blank answers. Pre and post covid were not defined by specific dates, so the answer to those specific survey questions could have been interpreted inconsistently. The survey itself took place over a 3-month period, implying that the number of personnel could have changed during the survey period itself.

## CONCLUSION

Our study findings demonstrate that overall lack of funding and staffing are commonly afflicting our rural health departments in the United States. Nurses were found to be the most utilized type of public health personnel, and health departments noted having adequate expertise despite having minimal professional experience and formal education in public health. There is a significant opportunity to utilize community collaboration to better health outcomes in rural communities.

### PUBLIC HEALTH IMPLICATIONS

The findings from this survey can be used to support rural public health departments when advocating for local policy, funding, and resources. These survey results illustrate the need for increased funding, and funding that can be flexibly used for different populations and issues specific to the area, public health personnel and infrastructure, assistance with transportation concerns, improved accessibility, mental/behavioral health services, and low-income clinics. These findings can also inform federal and state government efforts to enable rural areas to take fuller advantage of resources available and to inform future policy resource allocation. Additionally, our findings demonstrate potential opportunities for community engagement to fill gaps associated with traditional public health department structures.

## Data Availability

All data produced in the present study are available upon reasonable request to the authors

## Acknowledgements

We would like to acknowledge the championing of our survey by Tammy Moriearty at the Texas Department of State Health Services, everyone who took the time to answer our survey, and Rutgers School of Public Health at Rutgers University.

